# Body Composition and Body Image in Collegiate Athletes: The Mediating Role of Trait Sport Confidence

**DOI:** 10.64898/2026.04.24.26351348

**Authors:** Fengwei Hao, A. Mark Williams, Chengyi Liu, Sicong Liu

## Abstract

Athletes’ bodies are both performance agents and targets of evaluative scrutiny, yet little is known about the psychological processes linking body composition to body image among athletes. In this pre-registered study, we examined whether competence-related self-evaluations mediate or moderate associations between adiposity and body image in 327 Chinese collegiate athletes (78.6% male). Drawing on Self-Objectification Theory and the Sport Confidence Model, we tested two competing hypotheses, including a filter (parallel mediation) and a buffer (moderation) account. Factor analysis results of four body image scales supported a two-factor structure, comprising a proactive, functionality-oriented positive dimension and a reactive, appearance-distress-driven negative dimension.

Hierarchical regressions showed that fat mass index (FMI) was associated with lower positive and higher negative body image (*p*s < 0.05). Importantly, parallel mediation analysis results indicated that trait sport confidence mediated between FMI and both body image dimensions, with a stronger effect for positive body image (β = −0.04, 95% CI [−0.09, −0.01]) than for negative body image (β = 0.03, 95% CI [0.01, 0.07]). Subjective sport performance was not evidenced as a mediator. No moderation effects were supported. These findings suggest that the body composition–body image link in athletes is interpretive: enduring competence beliefs may matter more than proximal performance appraisals in affecting how athletes make sense of their bodies. Positive body image appears especially dependent on competence-grounded meaning-making, whereas negative body image remains more directly tied to appearance-based evaluative cues. Collegiate sport environments may benefit from prioritizing functionality-centered feedback over physique-focused evaluation.

## 1. Introduction

### 1.1 Body Image in Sports

Body image is a multidimensional psychological construct involving perceptions, evaluations, and behaviors related to the human body (Cash, 2004). Its research has been developed along two partially overlapping traditions. One tradition focuses on examining body image disturbance, particularly body dissatisfaction and eating pathology (Markey & Gillen, 2016; Quittkat et al., 2019), as well as the sociocultural processes through which concerns over appearance become normalized and psychologically consequential (Tiggemann, 2011). The other tradition centers on body competencies and physical self-perceptions, highlighting the body as a source of agency, function, and lived capability (Fox & Corbin, 1989; Franzoi, 1994). More recent work have advanced the understanding of body image by treating positive body image, including body appreciation and functionality appreciation, as a distinct construct rather than an absence of dissatisfaction (Aimé et al., 2020; Tort-Nasarre et al., 2023; Tylka & Wood-Barcalow, 2015). Accordingly, body appreciation refers to caring for the body and rejecting internalized sociocultural ideals (Avalos et al., 2005), and functionality appreciation highlights the significance of physical capabilities rather than aesthetic qualities (Alleva & Tylka, 2021).

The distinctions can be particularly meaningful in collegiate athletics, where the body is evaluated as a functional agent of performance as well as a visible object of scrutiny (Beckner & Record, 2016; Zaccagni & Gualdi-Russo, 2023). Athletes are therefore embedded within an environment in which performance demands, sport-specific body ideals, and broader sociocultural appearance norms are continuously evaluated. However, the form and intensity of these pressures may not be uniform across groups. These pressures may vary according to gender-specific appearance standards, the aesthetic and physiological demands of different sports, and the extent to which sporting competence becomes socially tied to bodily form. For female athletes, prior scholarship has long documented tensions between performance-related muscularity and culturally dominant ideals of femininity (Lundqvist et al., 2024). For male athletes, growing evidence suggests that body-related pressures increasingly center on muscularity, shaped by both sport-specific demands and broader masculine appearance norms (Galli et al., 2015; Steinfeldt et al., 2012). Across both contexts, the athletic body is not simply experienced; it is interpreted through overlapping lenses of competence, discipline, desirability, and social visibility. In this context, we examine how positive and negative body images are related but partially distinct dimensions rather than as opposite ends of a single continuum. Although appearance-based dissatisfaction remains closely linked to maladaptive body-related experiences, including body surveillance and social physique anxiety (Fitzsimmons-Craft et al., 2012; McCreary & Saucier, 2009), function-oriented body evaluations may support a more adaptive mode of self-relation by anchoring self-worth in embodied capability rather than external appearance. We examine these dimensions together to offer a more conceptually precise and ecologically grounded account of how collegiate athletes make sense of their bodies within performance-oriented yet appearance-focused environments.

### 1.2 Body Composition, Self-Objectification, and Sport Confidence

Within the evaluative climate of collegiate sport, body composition constitutes a salient input to body-related self-perception. In Self-Objectification Theory (SOT), Fredrickson and Roberts (1997) proposed that visible physical attributes (e.g., muscularity, adiposity) can increase perceived scrutiny and foster an internalized observer perspective. This process shifts attention from a first-person, embodied experience toward a third-person evaluative stance, increasing the likelihood that athletes view the body as an object to be judged rather than as a dynamic source of physical functionality (Kahalon et al., 2018).

Importantly, this objectification process does not unfold uniformly across sport settings.

In aesthetically judged sports, athletes may face explicit pressure to attain a particular physique because body form enters evaluative criteria (e.g., gymnastics). In other sports, bodily characteristics may be valued less for their appearance than for their perceived functional advantages, such as leanness in endurance-based events. In contrast, in some non-lean sports, greater body mass may be functionally valued, which may alter the form of body-related pressure (Chapa et al., 2022; Stoyel et al., 2019). Gender appearance norms may further intensify this tension for some female athletes, particularly when the body type valued for performance conflicts with broader feminine ideals (Reel et al., 2010; Soulliard et al., 2019).

From an SOT perspective, body composition is therefore unlikely to influence body image in a direct or uniform manner. Rather, the psychological consequences of body composition should depend on how athletes interpret their physical characteristics within the performance domain. That is, performance-related self-evaluations may serve as the cognitive mechanism through which objective physical markers acquire subjective body image significance. The Sport Confidence Model (SCM; Vealey, 1986; Vealey et al., 1998) offers a useful framework for understanding this interpretive process. SCM helps distinguish trait sport confidence, a relatively enduring belief about one’s athletic capability rooted in accumulated sport experiences, from state sport confidence, which is more momentary and responsive to recent performance feedback. More broadly, SCM suggests that competence-related appraisals shape how athletes make sense of performance demands and of themselves within sport settings (Vealey et al., 1998).

### 1.3 Indirect-Effect and Buffering Accounts

The integration of SOT and SCM suggests two competing explanatory accounts for how body composition may relate to body image. First, an indirect-effect/filter account proposes that associations between body composition and image may operate through performance-related self-evaluations. Favorable body composition may reinforce perceived athletic competence (trait sport confidence) and/or recent capability (subjective sport performance), supporting a more functionality-oriented interpretation of the body and, in turn, more positive body image. Conversely, perceived non-ideal body composition may undermine competence appraisals and increase susceptibility to an externalized observer perspective, elevating appearance-based concerns and body dissatisfaction. In this view, sport confidence and subjective performance may function as cognitive filters through which physical attributes acquire psychological meaning.

Alternatively, a moderation/buffer account proposes that performance-related self-evaluations serve as boundary conditions that determine the strength of the association between body composition and image. Specifically, high trait sport confidence or positive recent performance may buffer athletes from the negative implications of non-ideal body composition by anchoring self-worth in competence rather than appearance. In contrast, when confidence or perceived performance is low, objective physical attributes may become a more dominant, volatile determinant of body image. This account aligns with the idea that competence-based self-evaluations can protect mental health under scrutiny by shifting emphasis away from an externalized observer’s gaze and toward internalized functional capability.

### 1.4 The Present Study

Given these competing hypotheses, we examined the associations among body composition, performance-related self-evaluations, and multidimensional body image in collegiate athletes (pre-registered on AsPredicted: https://aspredicted.org/m4fb-36kq.pdf). Building on SOT and the SCM, we formulated two alternative models to clarify the psychological processes linking adiposity to body image (see Figure 1). Acknowledging an association between adiposity and body image (H1), the parallel mediation model (the filter account) includes that adiposity will show indirect associations with body image through trait sport confidence (H2 and H4) and state sport confidence (H3 and H5). This filter account assumes that, within a performance-oriented sport context, athletes may interpret their physical form through the lens of perceived athletic competence. Accordingly, higher adiposity may be associated with less positive and more negative body image primarily because it undermines confidence in one’s athletic ability. Alternatively, the moderation model (the buffer account) indicates that trait sport confidence (H6) and state sport confidence (H7) will moderate the association between adiposity and body image (H8). This buffer account suggests that higher levels of sport-related competence may protect athletes from the negative psychological implications of a less ideal body composition, thereby weakening the association between adiposity and body image.

**Figure 1.**
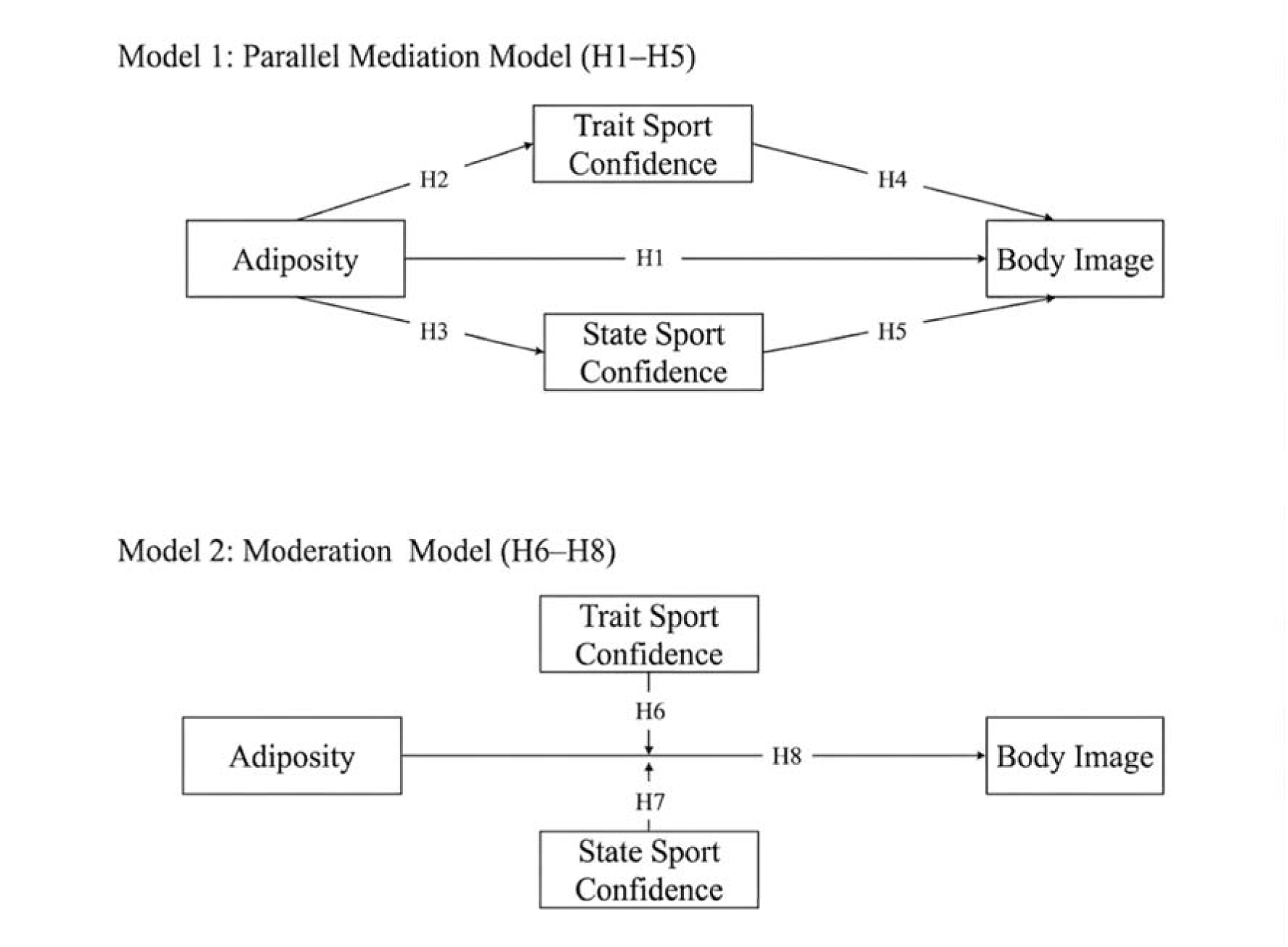
Conceptual models. H1 corresponds to Hypothesis 1, and this correspondence similarly applies to H2 through H8.

## 2. Methods

### 2.1 Participants and Procedures

A total of 370 local collegiate athletes participated. Individuals with medical conditions that compromised anthropometric assessments or influenced body-image evaluations, such as endocrine disorders, edema, or eating disorders, were excluded. Data were collected from two cohorts of freshmen entering college in the fall of 2024 and 2025, respectively.

Anthropometric assessments and psychological questionnaires were administered via a combined online–offline protocol supervised by trained research assistants. Ethical approval was obtained from the Ethics Committee of South China Normal University (Approval #: SCNU-SPT-2023-008). Participants were informed about the study purposes and procedures prior to providing informed consent.

### 2.2 Measures

#### Demographic information

Participants provided their demographic information, including gender, age, year in study, and sport type. Their competition levels were based on nationally recognized certification standards. Training load and history were assessed using variables such as weekly training frequency, per session duration, total weekly training time, and training age, with training intensity categorized into three levels according to the American College of Sports Medicine (ACSM) guidelines.

#### Body appreciation

Body appreciation was assessed using the Body Appreciation Scale–2 (BAS–2), a 10-item scale evaluating one’s acceptance, respect, and appreciation of one’s own bodies (Tylka & Wood-Barcalow, 2015). Items are rated on a 5-point Likert scale ranging from 1 (never) to 5 (always) with excellent observed consistency (Cronbach α = 0.93). We used the sum score with higher scores indicating greater body appreciation.

#### Functionality appreciation

Functionality appreciation was measured using the Functionality Appreciation Scale (FAS), which assesses one’s recognition and appreciation of body capabilities and functions (Alleva et al., 2017). Items are rated on a 5-point Likert scale ranging from 1 (never) to 5 (always) with excellent observed consistency (Cronbach α = 0.92). We used the sum score with higher values indicating greater appreciation of body functionality.

#### Body dissatisfaction

Body dissatisfaction was assessed using the Body Shape Questionnaire-Short Form (BSQ–8; Evans & Dolan, 1993). This 8-item scale helps evaluate cognitive and emotional concerns about body shape, with items rated on a 6-point Likert scale ranging from 1 (never) to 6 (always), while showing excellent observed internal consistency (Cronbach α = 0.92). We used the sum score with higher values indicating *higher* body dissatisfaction.

#### Weight bias internalization

Weight bias internalization was assessed using the 11-item Modified Weight Bias Internalization Scale (WBIS–M; Pearl & Puhl, 2014). Items are rated on a 7-point Likert scale ranging from 1 (strongly disagree) to 7 (strongly agree) with excellent observed consistency (Cronbach α = 0.88). We used the sum score with higher values indicating greater weight bias internalization.

#### Body composition

Height was measured using a stadiometer under standard procedures.

Body composition was assessed using the InBody 270 bioelectrical impedance analyzer according to the manufacturer’s standard procedures. Previous research has supported the reliability and validity of the InBody 270 bioelectrical impedance analyzer under standardized measurement conditions (Czartoryski et al., 2020; McLester et al., 2020). The measurement outputs included body mass index (BMI), FMI, and fat-free mass index (FFMI). FMI (kg/m²) was calculated as fat mass divided by height squared, and FFMI (kg/m²) was calculated as fat-free mass divided by height squared.

#### Sport confidence

Trait sport-confidence was assessed using the Trait Sport-Confidence Inventory (TSCI; Vealey, 1986), which includes 13 items to evaluate an athlete’s confidence about his/her sport-related abilities. Items are rated on a 9-point Likert scale ranging from 1 (low confidence) to 9 (high confidence) with excellent observed consistency (Cronbach α = 0.97). We used the sum score with higher values indicating greater confidence.

#### Sport performance

Subjective sport performance was assessed using the 2-item Subjective Performance Questionnaire (SPQ; Soulliard et al., 2019). Items are rated on an 11-point Likert scale ranging from 0 (extremely unsuccessful/unsatisfied) to 10 (extremely successful/satisfied) *with* excellent observed consistency (Cronbach α = 0.94). We used the sum score with higher values indicating greater self-perceived sport performance.

### 2.3 Analysis

We used the *lavaan* package in R with α level set at 0.05. During data preprocessing, we screened for potentially extreme observations using the 1.5 inter-quartile range (IQR) criterion and removed flagged cases after data checking. Univariate distributional properties were examined using skewness and kurtosis, and multivariate normality was evaluated using both Mardia’s tests and the Henze–Zirkler test. Because the data continued to show significant departures from multivariate normality, all subsequent confirmatory factor analyses (CFAs) were performed using robust maximum likelihood (MLR) estimation. This estimator was used to obtain more robust standard errors under non-normal conditions and was not intended to replace preprocessing decisions regarding extreme values. Table S1 in supplementary information shows descriptive statistics for all variables, including correlations among body composition measures, psychological scales, and relevant covariates, among which a strong polyserial correlation (*r* = 0.99) was identified between gender and FFMI, suggesting a high risk of collinearity. We therefore selected FFMI as the variable to represent information associated with gender (i.e., females have lower FFMI values) in all subsequent analyses.

To evaluate measurement properties, we conducted item-level CFAs on all 36 items from the BAS-2 (10 items), FAS (7 items), BSQ-8 (8 items), and WBIS-M (11 items). We specified two theory-informed measurement models and evaluated them using confirmatory factor analysis: a 2nd-order single-factor model, which treated all four scales as gauging one overarching latent construct, and a 2nd-order two-factor model, which specified that the scales loaded onto two distinct latent factors. We compared global model fit using multiple indices, including the chi-square statistic (χ^²^), the comparative fit index (CFI), the Tucker–Lewis index (TLI), the root mean square error of approximation (RMSEA), and the standardized root mean square residual (SRMR), according to the rule of thumb (Bentler, 1990; Hu & Bentler, 1999). When initial model fit was inadequate, limited model refinements were informed by modification indices and implemented sparingly to improve fit while minimizing overfitting. Once the superior model emerged, we extracted scores of the latent factor(s) and used such scores to represent body image.

To examine how body composition (i.e., FMI), gender, and sport type affect body image perception, we performed hierarchical regression analyses. We entered predictors in three nested models. Model 1 served as a baseline by merely including demographic and training-related variables (i.e., year in study, competition level, training intensity, weekly training duration, training age, and age). Model 2 went beyond Model 1 by further including FMI, gender, and sport type to test their main effects on body image. Model 3 extended Model 2 by considering FMI × Gender and FMI × Sport Type interactions, which helped test whether gender and/or sport type moderated the association between FMI and body image.

To answer the research question involving the two competing hypotheses, we performed two types of analysis. One type of analysis helped evaluate whether sport confidence (TSCI) and subjective sport performance (SPQ) mediate the association between FMI and body image. Specifically, we fit parallel mediation models using bias-corrected bootstrapping with 1,000 resamples, while controlling several covariates, including year in study, sport type, competition level, weekly training intensity and duration, training age, and age. The other type of analysis helped examine whether TSCI or SPQ moderate the association between FMI and body image. We performed analyses using regressions, and each model included the main effects of FMI and the moderator, the interaction term, and the same covariates used in the mediation models.

## 3. Results

### 3.1 Latent Factors of Body Image

Normality analysis results showed that the four scale scores (BAS, FAS, BSQ, and WBIS) contained no missing values. Although univariate skewness (-0.32 to 0.81) and kurtosis (2.33 to 3.08) were within acceptable ranges, Shapiro–Wilk tests for all four variables were significant (all *p* < 0.001), indicating departures from univariate normality. Multivariate normality was also not supported by the Henze–Zirkler test (HZ = 2.72, *p* < 0.001). Mardia’s tests confirmed this violation (skewness = 122.56, *p* < 0.001; kurtosis = 2.99, *p* = 0.003), supporting the use of the MLR estimator in the CFAs. Data preprocessing yielded a final sample of 327 collegiate athletes, including 257 males. Table 1 presents the demographic characteristics of the sample, and Table S1 in the Supplementary Information provides the correlation matrix for all observed variables. Notably, the four body image measures appeared to cluster into two distinct subdomains: one comprising BAS and FAS, and the other comprising BSQ and WBIS. Specifically, all correlations within subdomains were positive (*r*_BAS-FAS_ = 0.71, *r*_BSQ-WBIS_ = 0.51) and those between subdomains were negative (*r*_BAS-BSQ_ = −0.20, *r*_BAS-WBIS_ = −0.17, *r*_FAS-BSQ_ = −0.02, *r*_FAS-WBIS_ = −0.15), an observation that was consistent with the two-component model of body image.

**Table 1.** Sample demographic information.

| Sample Characteristic | % (n) or M (SD) |
| --- | --- |
| Gender<sup>a</sup> |  |
| Male | 78.59 (257) |
| Female | 21.41 (70) |
| Year in Study<sup>a</sup> |  |
| 2024 | 30.58 (100) |
| 2025 | 69.42 (227) |
| Sport Type<sup>a</sup> |  |
| Non-aesthetic/Non-lean | 76.54 (251) |
| Aesthetic/Lean | 23.46 (76) |
| Competition Level<sup>a</sup> |  |
| Elite | 3.06 (10) |
| First Class | 69.11 (226) |
| Second Class | 23.55 (77) |
| Third Class | 4.28 (14) |
| Training Intensity<sup>a</sup> |  |
| High | 24.77 (81) |
| Moderate | 70.03 (229) |
| Low | 5.20 (17) |
| Weekly Frequency of Athletic Training (sessions/week)<sup>a</sup> | 4.52 (1.32) |
| Training Duration per Session (min/sessions)<sup>a</sup> | 103.15 (39.97) |
| Training Duration per Week (min/week)<sup>a</sup> | 478.06 (253.51) |
| Training Age (month)<sup>a</sup> | 34.58 (42.53) |
| Age (year)<sup>a</sup> | 20.14 (1.37) |
| Height (cm)<sup>a</sup> | 175.21 (7.20) |
| Weight (kg)<sup>a</sup> | 68.57 (8.89) |
| Body Mass Index (kg/m<sup>2</sup>)<sup>a</sup> | 22.30 (2.24) |
| Fat-Free Mass Index (kg/m<sup>2</sup>)<sup>a</sup> | 16.82 (1.84) |
| Fat Mass Index (kg/m<sup>2</sup>)<sup>a</sup> | 3.00 (1.25) |

*Note.* <sup>a</sup> dichotomous variable. <sup>b</sup> ordinal variable. <sup>c</sup> continuous variable. Dichotomous and ordinal variables are summarized using percentage and count, % (n), and continuous variables using mean and standard deviation, M (SD). Year in Study = year in which the questionnaire was administered; Competition Level = certified level of sport competition; Training Intensity = self-reported volume of athletic training classified based on the American College of Sports Medicine and the American Heart Association standards; Weekly Frequency of Athletic Training = average number of training sessions per week; Training Duration per Session = duration of a single athletic training session; Training Duration per Week = total duration spent on athletic training over a week; Training Age = total number of month receiving athletic training; Age = year of age.

We conducted item-level CFAs to compare the two competing higher-order measurement models. The one-factor higher-order model showed inadequate fit to the data, χ²(590) = 1334.48, CFI = 0.88, TLI = 0.87, RMSEA = 0.07, SRMR = 0.12. Guided by the modification indices, we revised the model to freely estimate one residual covariance between two WBIS items. Although this adjustment produced slight improvement, the overall fit remained below acceptable levels, χ²(589) = 1279.96, CFI = 0.89, TLI = 0.88, RMSEA = 0.07, SRMR = 0.12.

We next specified a two-factor higher-order model, which demonstrated better overall fit than the one-factor model, χ²(589) = 1237.07, CFI = 0.90, TLI = 0.89, RMSEA = 0.06, SRMR = 0.06. After freely estimating the same residual covariance between the two WBIS items, model fit improved further, χ²(588) = 1180.02, CFI = 0.91, TLI = 0.90, RMSEA = 0.06, SRMR = 0.06. Table S2 in the Supplementary Information summarizes the fit indices for all tested measurement models. We therefore retained the revised two-factor higher-order model (i.e., 2-Factor + WBISpath in Table S2 in the supplementary information) as the final measurement model and extracted latent factor scores for subsequent analyses. Figure 2 displays the model, featuring a positive component of body image (i.e., positive BI) and a negative component of body image (i.e., negative BI).

**Figure 2.**
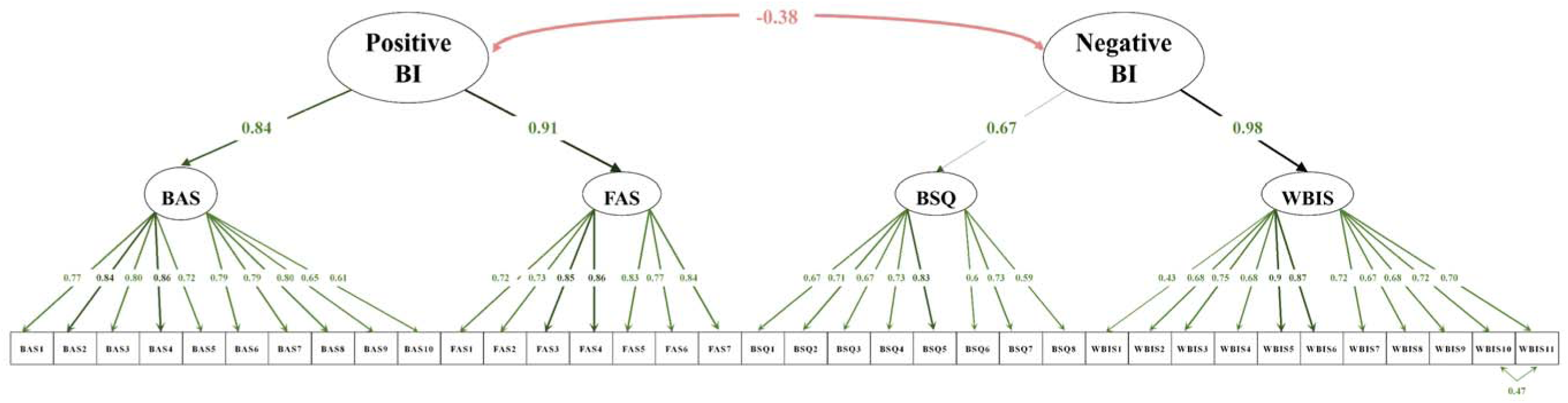
High order measurement model of body image components. BI = body image; BAS_Factor = latent factor extracted from items of the body appreciation scale; FAS_Factor = latent factor extracted from items of functionality appreciation scale; BSQ_Factor = latent factor extracted from items of the body shape questionnaire; WBIS_Factor = latent factor extracted from items of the weight bias internalization scale.

### 3.2 Effect of Body Composition, Gender, and Sport Type on Body Image

Table 2 shows results of hierarchical regression analyses for both positive and negative components of body image. For positive body image, moderate training intensity was negatively associated with positive body image and remained significant across Models 1–3 (*ps* < 0.03). After adding the focal predictors in Model 2, FMI was negatively associated with positive body image, *B* =-0.10, *t*(*314*) =-2.08, *p* < 0.05, but no interactions were identified (*ps* > 0.73) in Model 3. For negative body image, age was negatively associated with negative body image and remained significant across Models 1–3 (*ps <* 0.04). In Model 2, FMI showed a significant positive association with negative body image, *B* = 0.22, *t*(*314*) = 4.78, *p* < 0.001. In Model 3, no significant interactions were observed (*ps* > 0.41). Taken together, these findings suggest that higher FMI was associated with lower positive body image and higher negative body image, whereas neither gender nor sport type was significantly associated with either dimension.

**Table 2.**
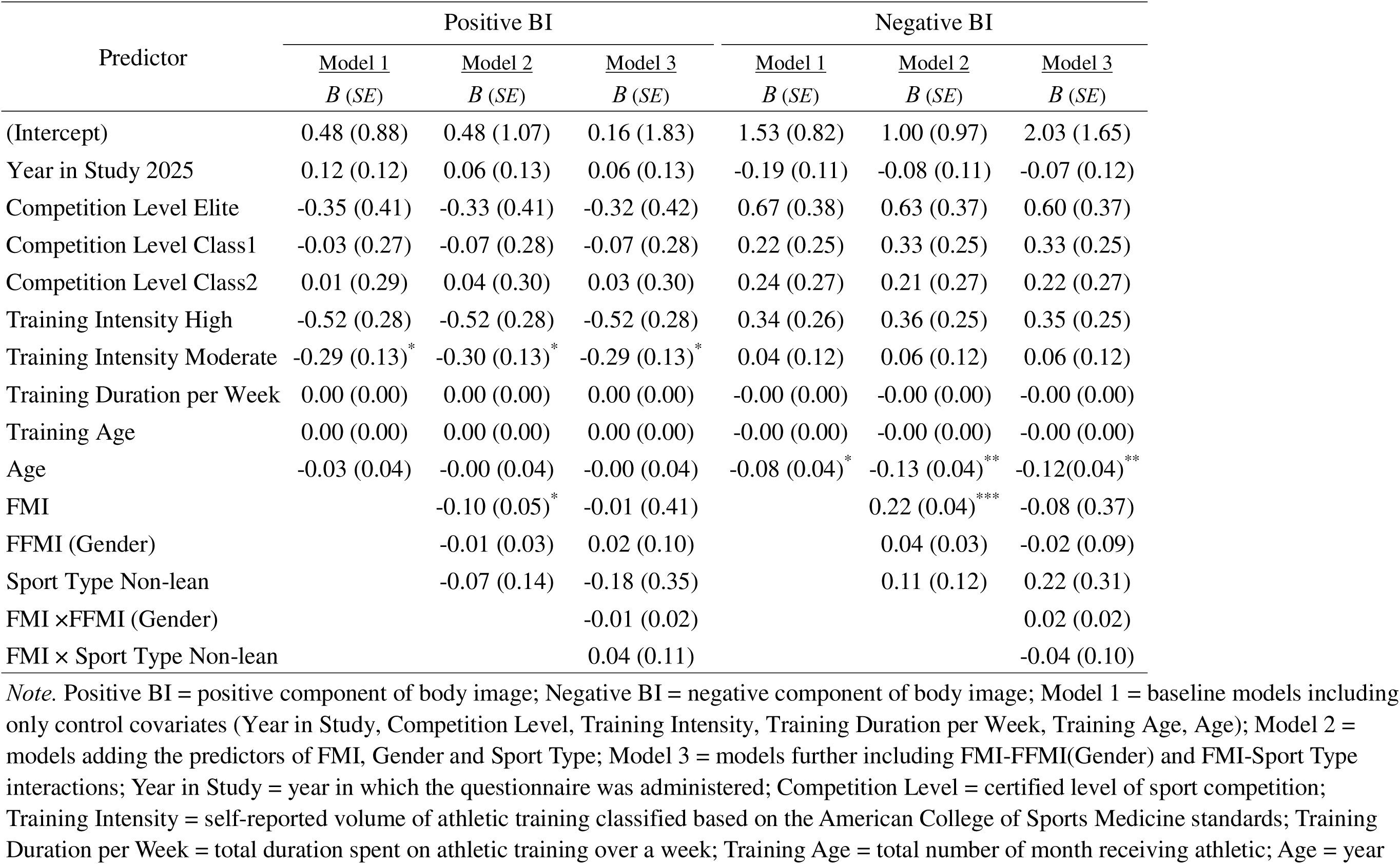
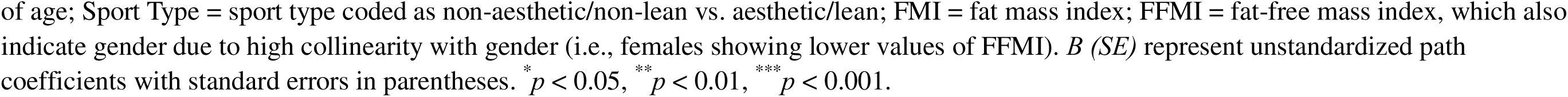
Regression Analyses Predicting Positive and Negative Body Image.

| Predictor | Positive BI |  |  | Negative BI |  |  |
| --- | --- | --- | --- | --- | --- | --- |
|  | <u>Model 1</u> | <u>Model 2</u> | <u>Model 3</u> | <u>Model 1</u> | <u>Model 2</u> | <u>Model 3</u> |
|  | *B (SE)* | *B (SE)* | *B (SE)* | *B (SE)* | *B (SE)* | *B (SE)* |
| (Intercept) | 0.48 (0.88) | 0.48 (1.07) | 0.16 (1.83) | 1.53 (0.82) | 1.00 (0.97) | 2.03 (1.65) |
| Year in Study 2025 | 0.12 (0.12) | 0.06 (0.13) | 0.06 (0.13) | -0.19 (0.11) | -0.08 (0.11) | -0.07 (0.12) |
| Competition Level Elite | -0.35 (0.41) | -0.33 (0.41) | -0.32 (0.42) | 0.67 (0.38) | 0.63 (0.37) | 0.60 (0.37) |
| Competition Level Class1 | -0.03 (0.27) | -0.07 (0.28) | -0.07 (0.28) | 0.22 (0.25) | 0.33 (0.25) | 0.33 (0.25) |
| Competition Level Class2 | 0.01 (0.29) | 0.04 (0.30) | 0.03 (0.30) | 0.24 (0.27) | 0.21 (0.27) | 0.22 (0.27) |
| Training Intensity High | -0.52 (0.28) | -0.52 (0.28) | -0.52 (0.28) | 0.34 (0.26) | 0.36 (0.25) | 0.35 (0.25) |
| Training Intensity Moderate | -0.29 (0.13)* | -0.30 (0.13)* | -0.29 (0.13)* | 0.04 (0.12) | 0.06 (0.12) | 0.06 (0.12) |
| Training Duration per Week | 0.00 (0.00) | 0.00 (0.00) | 0.00 (0.00) | -0.00 (0.00) | -0.00 (0.00) | -0.00 (0.00) |
| Training Age | 0.00 (0.00) | 0.00 (0.00) | 0.00 (0.00) | -0.00 (0.00) | -0.00 (0.00) | -0.00 (0.00) |
| Age | -0.03 (0.04) | -0.00 (0.04) | -0.00 (0.04) | -0.08 (0.04)* | -0.13 (0.04)** | -0.12(0.04)** |
| FMI |  | -0.10 (0.05)* | -0.01 (0.41) |  | 0.22 (0.04)*** | -0.08 (0.37) |
| FFMI (Gender) |  | -0.01 (0.03) | 0.02 (0.10) |  | 0.04 (0.03) | -0.02 (0.09) |
| Sport Type Non-lean |  | -0.07 (0.14) | -0.18 (0.35) |  | 0.11 (0.12) | 0.22 (0.31) |
| FMI ×FFMI (Gender) |  |  | -0.01 (0.02) |  |  | 0.02 (0.02) |
| FMI × Sport Type Non-lean |  |  | 0.04 (0.11) |  |  | -0.04 (0.10) |

*Note.* Positive BI = positive component of body image; Negative BI = negative component of body image; Model 1 = baseline models including only control covariates (Year in Study, Competition Level, Training Intensity, Training Duration per Week, Training Age, Age); Model 2 = models adding the predictors of FMI, Gender and Sport Type; Model 3 = models further including FMI-FFMI(Gender) and FMI-Sport Type interactions; Year in Study = year in which the questionnaire was administered; Competition Level = certified level of sport competition; Training Intensity = self-reported volume of athletic training classified based on the American College of Sports Medicine standards; Training Duration per Week = total duration spent on athletic training over a week; Training Age = total number of month receiving athletic; Age = year
of age; Sport Type = sport type coded as non-aesthetic/non-lean vs. aesthetic/lean; FMI = fat mass index; FFMI = fat-free mass index, which also indicate gender due to high collinearity with gender (i.e., females showing lower values of FFMI). *B (SE)* represent unstandardized path coefficients with standard errors in parentheses. \* $p < 0.05$ , \*\* $p < 0.01$ , \*\*\* $p < 0.001$ .

**Table 3.**
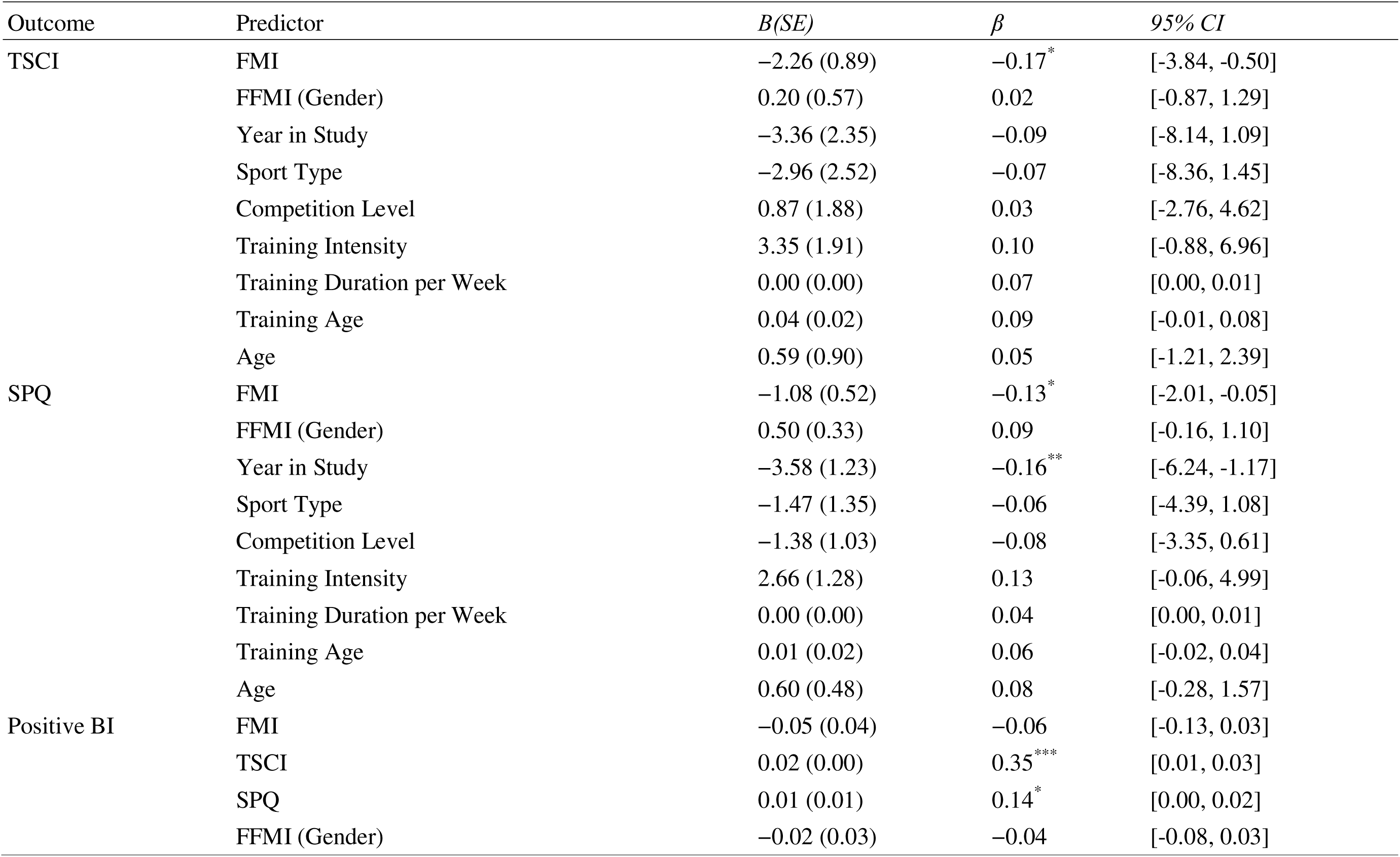
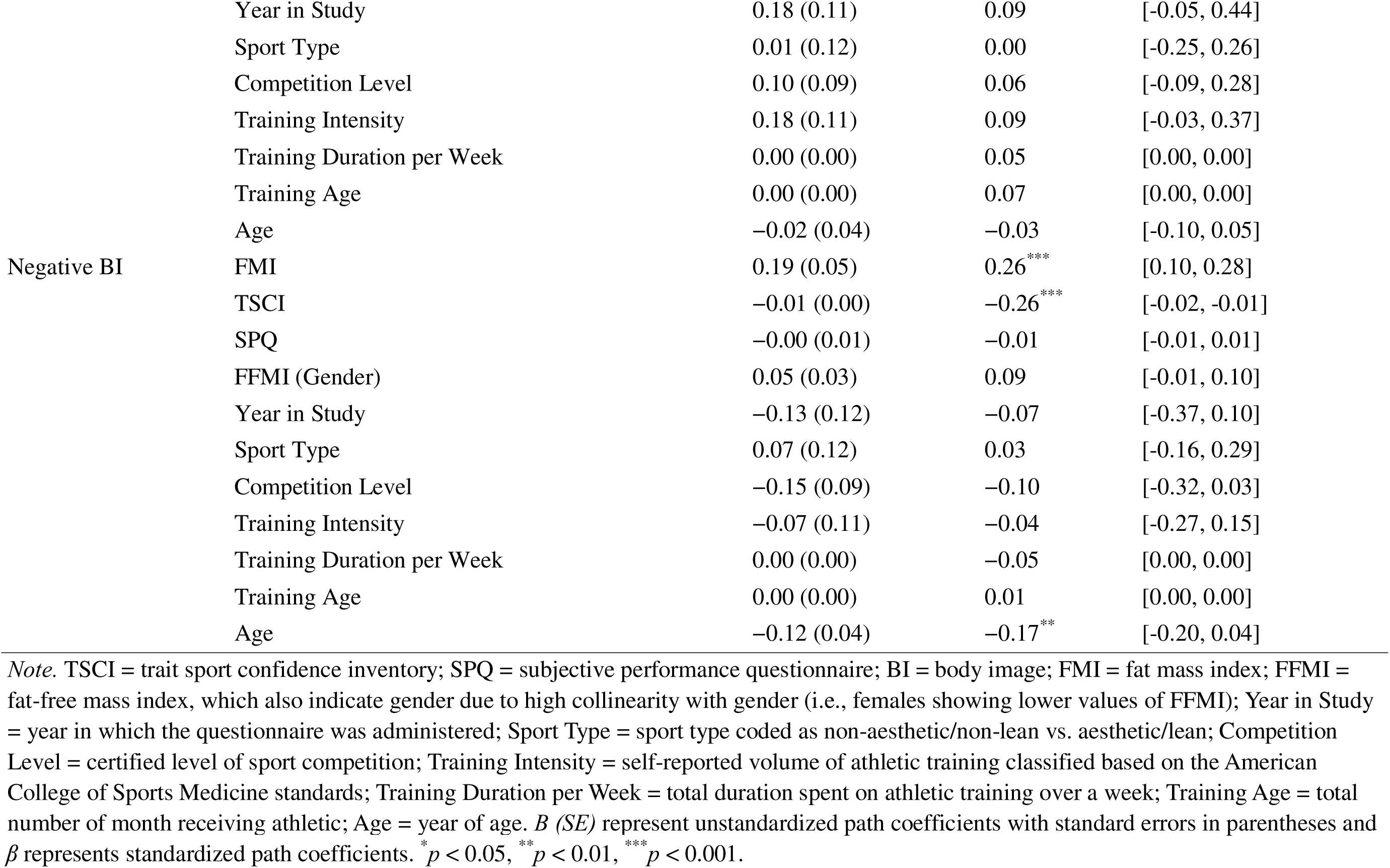
Standardized Path Coefficients for Parallel Mediation Model with FMI.

### 3.3 Roles of Sport Confidence and Subjective Sport Performance

#### Parallel Mediation Analysis

Figure 3a displays results from testing TSCI and SPQ as two parallel mediators between FMI and positive body image. Specifically, in presence of an indirect effect through TSCI (β = −0.04, 95% CI [−0.09, −0.01]), neither a direct effect of FMI (β = −0.06, 95% CI [−0.13, 0.03]) nor an indirect effect through SPQ (β = −0.01, 95% CI [−0.04, 0.00]) was identified. Figure 3b illustrates results from testing TSCI and SPQ as two parallel mediators between FMI and negative body image. Unlike results for the positive BI in Figure 3a, the direct effect of FMI remained significant (β = 0.26, 95% CI [0.10, 0.28]) in the presence of a significant indirect effect of TSCI (β = 0.03, 95% CI [0.01, 0.07]) and a non-significant indirect effect of SPQ (β =-0.00, 95% CI [−0.02, 0.02]).

**Figure 3.**
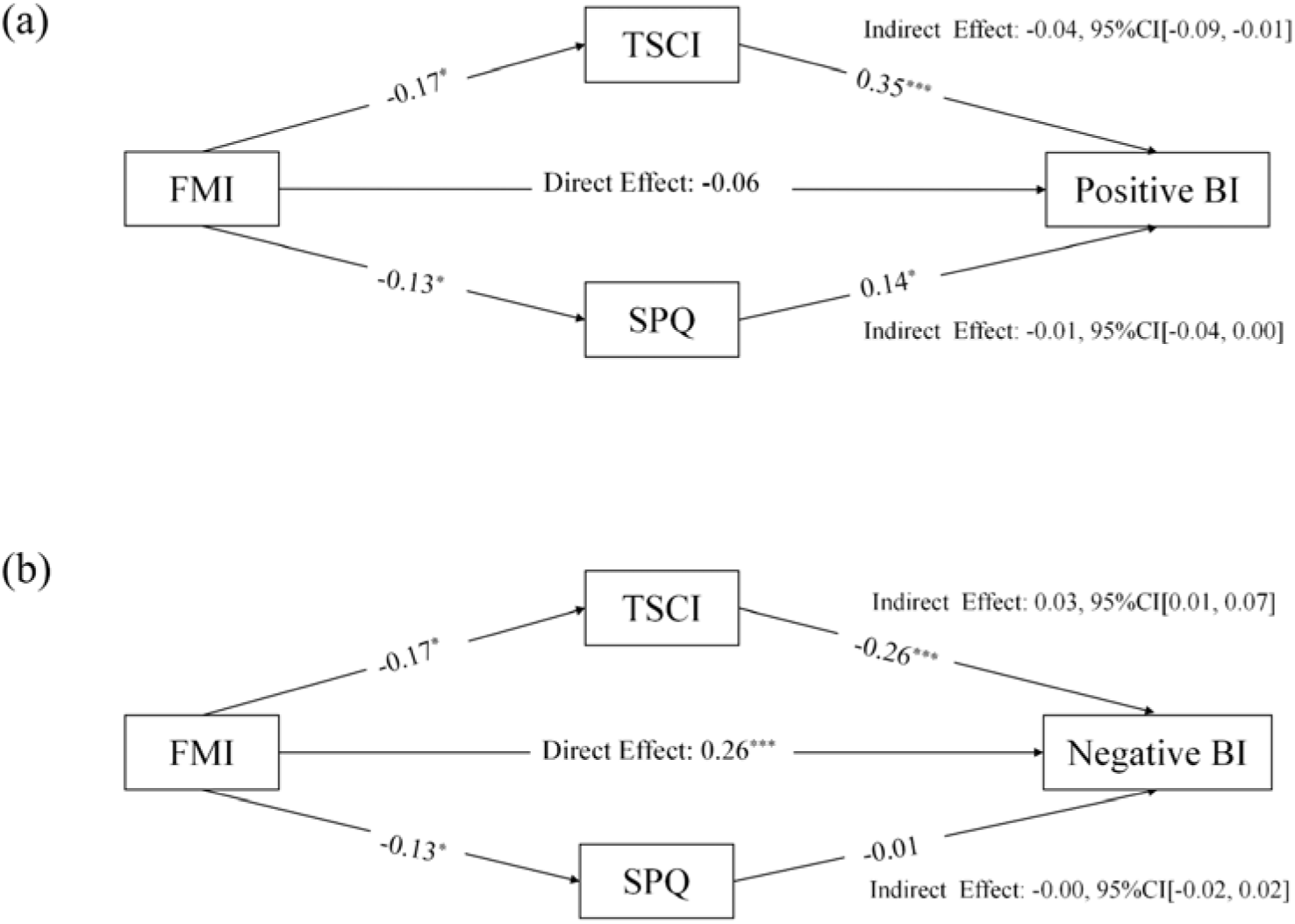
Parallel mediation model results. Panel (a) illustrates the parallel mediation model in which the association between FMI and positive body image was statistically accounted for by TSCI, while the indirect effect of SPQ was non-significant. Panel (b) displays the parallel mediation model in which the association between FMI and negative body image was partially statistically accounted for by TSCI. The effects of FFMI, TSCI, SPQ, Year in Study, Sport Type, Competition Level, Training Intensity, Training Duration per Week, Training Age, and Age were controlled for at each endogenous variable. FFMI = fat-free mass index; Year in Study = year in which the questionnaire was administered; Sport Type = sport type coded as non-aesthetic /non-lean vs. aesthetic/lean; Competition Level = certified level of sport competitiveness; Training Intensity = self-reported volume of athletic training classified based on the American College of Sports Medicine and the American Heart Association standards; Training Duration per Week = total duration spent on athletic training over a week; Training Age = total number of month receiving athletic training; Age = year of age; FMI = fat mass Index; TSCI = trait sport confidence inventory; SPQ = subjective performance questionnaire; BI = body image. \**p* < 0.05, \*\**p* < 0.01, \*\*\**p* < 0.001.

#### Moderation Analysis

We identified no interactions (i.e., FMI *×* TSCI, FMI *×* SPQ) on either positive body image or negative body image (*p*s > 0.16), although being older in age was associated with lower negative body image (*p*s < 0.001). Table S3 in the Supplement Information has details. The results were more consistent with the hypothesized indirect-effect pattern than with the moderation model. The current sample did not provide statistical support for moderation by sport confidence or subjective sport performance, though the limited power to detect interaction effects in observational designs (McClelland & Judd, 1993) means these null findings should be interpreted with caution.

## 4. Discussion

This present study helps examine the psychological mechanisms through which objective body composition is associated with multidimensional body image in Chinese collegiate athletes. Drawing on Self-Objectification Theory and the Sport Confidence Model, we evaluated two competing explanatory accounts: a filter account, in which performance-related self-evaluations mediate the association between adiposity and body image, and a buffer account, in which such evaluations moderate that association. Our findings were more consistent with the filter than the buffer account. Trait sport confidence emerged as a significant mediator of the association between FMI and both positive and negative body image, whereas neither TSCI nor SPQ moderated these associations. The following sections interpret these results in turn and situate them within the broader theoretical and applied literature.

### 4.1 A Two-Factor Structure of Body Image in Collegiate Athletes

The measurement results supported a two-factor higher-order representation of body image that distinguished a positive component (grounded in body appreciation and functionality appreciation) from a negative component (indexed by body dissatisfaction and weight bias internalization). This two-factor solution provided meaningfully better fit than a unidimensional alternative, consistent with theoretical arguments that positive body image is not merely the low end of a dissatisfaction continuum but rather involves partially distinct psychological processes (Tylka & Wood-Barcalow, 2015). In collegiate sports, this distinction may reflect the fact that athletes routinely evaluate their bodies both as agents of performance and as objects of social scrutiny (Beckner & Record, 2016; Zaccagni & Gualdi-Russo, 2023), suggesting that appreciation-oriented and distress-oriented body evaluations may be differentially responsive to the demands and pressures that characterize this environment.

The positive factor, which integrated body appreciation and functionality appreciation, reflects an orientation toward what the body can accomplish rather than how it appears. It represents a mode of embodied engagement that has been linked to adaptive psychological outcomes across general and athlete populations (Aimé et al., 2020; Alleva & Tylka, 2021). The negative factor, which captured appearance-based distress and internalized weight stigma, reflects a self-evaluative stance directed primarily at how the body looks relative to social standards of acceptability, which closely parallels the objectified self-perception described in SOT (Fredrickson & Roberts, 1997). The differential associations with FMI and sport confidence observed across factors, described in subsequent sections, underscore the value of treating these two dimensions as conceptually and empirically separable outcomes in research on athlete body image.

### 4.2 Body Composition and the Multidimensionality of Body Image

We found that FMI was consistently associated with both positive and negative body image, suggesting that adiposity was related to body image in a pattern broadly consistent with two distinct theoretical perspectives. From the standpoint of SOT, higher FMI may be associated with greater appearance-based self-evaluation, which may correspond to elevated body-related distress. From the perspective of the SCM, FMI may be viewed in relation to perceptions of physical readiness, sharing variance with the competence-based evaluations relevant to body appreciation. Thus, rather than indicating a directly observed psychological process, the present findings suggest a pattern of associations that is compatible with both appearance-focused and competence-focused interpretations of body image. These associations did not differ significantly by gender or sport type in this sample. Although we used the conventional lean/aesthetic versus non-lean/non-aesthetic classification to facilitate comparability with prior work (Burgon et al., 2023), the absence of moderation suggests that physique-and performance-related evaluative pressures may be pervasive across contemporary collegiate sport settings. Under such conditions, FMI may remain a salient reference point in athletes’ body-related self-evaluations even when sport-specific demands differ.

### 4.3 Trait Sport Confidence as an Interpretive Pathway

The association between FMI and both body image dimensions was mediated by TSCI in that higher FMI was associated with lower TSCI, which in turn was associated with lower positive and higher negative body image. In the presence of this indirect effect, the direct association between FMI and positive body image was fully attenuated, whereas for negative body image a significant direct association remained alongside the indirect effect. The asymmetry in indirect effect size across dimensions is theoretically informative. Positive body image, as characterized by functionality appreciation and bodily respect, may be especially dependent on competence-grounded meaning-making because its core content, valuing what the body can do, is directly instantiated by sport confidence. However, negative body image is driven by appearance-focused distress and weight stigma, which may be more directly elicited by objective physical characteristics through an appearance-evaluation pathway that operates partly independently of sport confidence. Such an interpretation integrates two theoretical frameworks in that the SCM supplies a competence-based lens through which physical characteristics acquire body image significance, and SOT accounts for the appearance-evaluation pathway that more directly drives body dissatisfaction. The mediating role of TSCI is also consistent with recent athlete evidence linking sport confidence to positive body image facets (Ricketts et al., 2023; Soulliard et al., 2019) and extends this work by situating sport confidence within a body composition–body image relationship.

Unlike TSCI, SPQ did not demonstrate a comparable mediating role. This null result carries theoretical implications in that SPQ captures athletes’ proximal, context-sensitive appraisals of recent performance, which may fluctuate in response to competitive outcomes, training load, and coach feedback. Considering that TSCI reflects a more stable and generalized belief about athletic capability accumulated across repeated sport experiences (Vealey, 1986), the present results suggest that it is this stable, cross-contextual sense of competence, rather than satisfaction with recent performance, that functions as the primary cognitive bridge between body composition and body image.

### 4.4 Absence of Buffering Effects

Neither TSCI nor SPQ moderated the FMI–body image association, providing no support for the buffer account. Prior to interpreting these null results substantively, it is worth noting that interaction effects in observational designs are typically small and difficult to detect, particularly given the sample sizes common in athlete research (Aguinis et al., 2005; McClelland & Judd, 1993). Beyond statistical power, there are also theoretical reasons to expect limited buffering in competitive collegiate contexts: body composition indices are routinely treated as performance-relevant information in these environments, and even athletes with high sport confidence may find it difficult to cognitively discount their body composition when it is continuously reinforced as meaningful by training practices and normative standards. Therefore, buffering effects may be more likely to emerge in less evaluatively intense contexts or following interventions that specifically decouple body composition evaluation from athletic identity.

### 4.5 Practical Implications

The findings carry practical implications for collegiate sport environments. Because FMI was linked to positive body image partly through its association with sport confidence, approaches that support competence-based self-appraisal (e.g., mastery-oriented feedback, skill-referenced goal-setting, evaluative climates that minimize social comparison) may be more effective for promoting positive body image than approaches focused on appearance or body composition norms alone. At the same time, the persistence of a direct effect of FMI on negative body image indicates that confidence-building alone is unlikely to fully offset appearance-based distress. Broader environmental modifications, such as limiting unsolicited body composition feedback, protecting the confidentiality of anthropometric data, and reducing physique-focused evaluation, are likely necessary complements. These dual emphases, on building competence beliefs and restructuring evaluative conditions, are probably more effective in combination than either approach individually.

### 4.6 Limitations and Future Directions

Several limitations warrant consideration. First, the cross-sectional design precludes causal inference. Although the indirect role of TSCI is theoretically grounded and statistically consistent with mediation, reverse-causal and third-variable accounts cannot be excluded.

Future research should adopt longitudinal designs, such as cross-lagged panel models or experience sampling, to establish the temporal ordering of body composition, sport confidence, and body image across athletes’ collegiate development. Second, the male-majority sample (78.6%) raises questions about the generalizability of SOT-derived interpretations. The original theory centers on the sexual objectification of women, and its application to male athletes, for whom scrutiny may center more on performance capacity or muscularity, may require recalibration, particularly in Chinese cultural contexts where traditional masculinity norms shape the meaning of athletic physicality. Future work should test measurement invariance across gender and explore whether the mediating role of TSCI operates equivalently for female and male athletes. Third, FMI captures only one facet of the physical self; muscularity, power capacity, and body proportionality may operate through distinct pathways and should be incorporated in future research. Finally, the lean versus non-lean sport classification is a coarse proxy for the evaluative ecology athletes inhabit; proximal environmental indicators (e.g., coach feedback practices, body composition monitoring policies, peer norms) would more precisely capture the contextual conditions under which adiposity acquires body image significance. Replication in culturally diverse samples is also needed to test whether the present pathways generalize beyond Chinese collegiate sport.

### 4.7 Conclusions

This study identifies trait sport confidence as a significant interpretive pathway through which body composition is associated with multidimensional body image in collegiate athletes. The mediating role of TSCI was stronger for positive than for negative body image, suggesting that functionality-oriented body appreciation is especially dependent on competence-grounded self-evaluation, whereas appearance-based distress retains a more direct relationship with adiposity. The absence of buffering effects suggests that confidence alone may not offset the psychological significance of body composition in evaluatively intensive sport environments. Collectively, these findings support dual-focus approaches that both cultivate competence-based body meaning-making and reduce appearance-centered evaluative pressures in collegiate sport settings.

## Contributors

Conceptualization and design: all authors

Data acquisition, analysis, or interpretation: FH SL Drafting of the manuscript: SL FH AMV

Funding obtainment: CL SL

## Data Sharing Statement

No additional data are available.

## Declaration of interests

None reported.

## Funding/Support

None Funding.

## Data Availability

All data produced in the present study are available upon reasonable request to the authors

## Acknowledgement

We sincerely thank Dr. Robert C. Eklund for his feedback on an earlier version of this manuscript.

## Supplementary Material (1)

Supplementary appendix

## Notes

### Competing Interest Statement

The authors have declared no competing interest.

### Funding Statement

This study did not receive any funding

### Author Declarations

Ethical approval was obtained from the Ethics Committee of South China Normal University (Approval #: SCNU-SPT-2023-008).

